# LynchRisk: a Bayesian network to estimate the probability of Lynch syndrome based on pedigree and tumor characteristics

**DOI:** 10.64898/2026.10.14.26365520

**Authors:** Alexandra Lefebvre, Patrick Benusiglio, Grégory Nuel

## Abstract

**Background:** Current Lynch syndrome (LS) prediction models do not account for detailed family structure and tumour mismatch repair (MMR) status. We developed LynchRisk, a Bayesian model that estimates individualized LS probability from pedigree, cancer history and tumour MMR test results.

**Methods:** LynchRisk combines Mendelian inheritance, age- and sex-specific cancer hazards, affected and unaffected relatives, and detailed MMR status. Parameters were derived from the literature.

**Results:** In two simulated cases of probands with LS-associated cancers, family structure and MMR status markedly changed LS probability estimates, with implications for germline testing decisions. LynchRisk also identified situations in which a negative germline result might be considered sufficient to rule out LS or, conversely, prompt further investigations such as somatic MMR or long-read sequencing.

**Discussion:** LynchRisk gives individualized LS probability by combining clinical, family, and tumour data at different stages of the diagnostic process. Further work will focus on recalibrating cancer hazard functions, validating model performance through simulations, and developing an R package for use by interested research groups.

## Introduction

Universal germline testing in patients with Lynch syndrome (LS) associated cancers, albeit explored in specific clinical or research settings, is unlikely to be widely implemented internationally ^1;2^. The reasons are the associated costs, the inability of laboratories – especially academic ones - to carry out testing on such a large scale, and the lack of trained professionals ^3^. Patient selection is therefore likely to remain the rule in the medium term^4;5^. Models estimating hereditary cancer syndrome (HCS) probability have been developed to assist with patient selection, with thresholds guiding the testing decision. CanRisk is a widely endorsed tool that estimates hereditary breast and ovarian cancer probabilities ^6^. In LS, the best-known models are PREMM5 and its pan-HCS iteration PREMMplus ^7^. Both show satisfactory performances, with areas under the curve (AUC) ranging from 0.68 to 0.94, the highest AUC being for PREMM5 and LS limited to MLH1/MSH2/EPCAM.

LS-associated cancers have a mismatch repair-deficient phenotype (MMRd) that differentiates them from the majority of non-LS cancers. As a result, the MMR status is a major parameter in germline testing decisions. For example, 55% of MMRd endometrial cancers without MLH1 promoter hypermethylation are caused by LS^8^. On the contrary, MMR protein expression makes LS unlikely given its >90-95% negative predictive value ^9;10^. A limitation of both PREMM5 and PREMMplus is that neither account for the MMRd / MMR-proficient (MMRp) status. LS probability is undoubtedly much different in a 45-year-old patient with MMRd vs. MMRp cancer and no cancer family history (FH). Incorporating MMR status could therefore improve LS probability estimation. Furthermore, the models only integrate family members with a LS-associated cancer history, and not unaffected ones. In a large family, a high number of relatives free of colorectal, endometrial and urothelial cancer is most likely a negative predictor of LS.

To address these limitations, we have developed LynchRisk, an experimental Bayesian model that integrates personal cancer history, MMRd/MMRp status, LS-associated cancer FH and family structure within a unified probabilistic framework. LynchRisk aims to provide more accurate LS probability estimates in order to better support germline testing decisions.

## Methods

LynchRisk is a generative Bayesian model of Lynch syndrome that combines Mendelian inheritance, age- and sex-specific cancer hazard functions, FH including affected and unaffected relatives, personal cancer history and tumor characteristics including somatic IHC (assessing MLH1/PMS2 and MSH2/MSH6 dimers, as well as isolated MSH6 and PMS2), MSI testing, MLH1 promoter hypermethylation analysis, and BRAF V600E mutation testing.

It considers up to two cancers per person in six cancer sites: colon, rectum, endometrium, ovary, upper gastrointestinal tract (which aggregates stomach, small bowel, bile duct, gall-bladder, and pancreas) and urinary tract (which aggregates ureter, renal pelvis, and urinary bladder). Latent genotypes are assumed to be composed of zero, one or two PV(s) in different MRR genes.

### Parameter

We use piecewise constant hazard functions with age intervals of 5 years from 25 to 75 years old. Incidence rates are calibrated from cancer incidence rates in the French general population estimated by Defossez et al. ^11^ and incidence data in LS carriers estimated by Dominguez-Valentin et al. ^12^ over the Prospective Lynch Syndrome Database (PLSD, https://plsd.eu) (see Appendix A for details on the calibration process along with Tables 1 and 2 for calibrated incidence rates, per 100,000 person-years, for males and females respectively). Moreover, we assume an additive effect of MMR PVs on the hazard scale for carriers of two PVs (in different MMR genes). Finally, hazard functions are considered independent from the past history of disease ^13^.

**Table 1:** Incidence rate estimates in males per 100,000 person-years. Columns represent age intervals, rows represent cancer sites, and row groups represent non-carriers and carriers of a single pathogenic variant in MLH1, MSH2, MSH6, or PMS2. Upper GI aggregates stomach, small bowel, bile duct, gallbladder, and pancreas. Urinary tract aggregates ureter, renal pelvis, and urinary bladder.

|  | 0-25 | 25-30 | 30-35 | 35-40 | 40-45 | 45-50 | 50-55 | 55-60 | 60-65 | 65-70 | 70-75 | 75-+ |
| --- | --- | --- | --- | --- | --- | --- | --- | --- | --- | --- | --- | --- |
| **Non-Carrier** |  |  |  |  |  |  |  |  |  |  |  |  |
| Colon | 0.7 | 1.8 | 2.6 | 3.6 | 5.9 | 11.1 | 24.2 | 45.9 | 78.2 | 116.2 | 157.8 | 210.3 |
| Rectum | 0.3 | 0.6 | 1.4 | 2.3 | 4.6 | 10.5 | 22.8 | 37.5 | 54.8 | 76.3 | 94.1 | 107.3 |
| Upper GI | 0.6 | 1.1 | 2.0 | 4.2 | 9.0 | 18.3 | 34.2 | 55.5 | 84.3 | 122.6 | 162.3 | 201.0 |
| Urinary tract | 0.6 | 1.4 | 3.9 | 8.0 | 14.7 | 25.9 | 46.9 | 83.0 | 131.7 | 182.8 | 233.8 | 298.8 |
| **MLH1** |  |  |  |  |  |  |  |  |  |  |  |  |
| Colon | 0.7 | 570.3 | 803.6 | 1114.7 | 1814.5 | 3369.9 | 1390.8 | 2570.5 | 1062.7 | 1600.1 | 2168.1 | 2880.1 |
| Rectum | 0.3 | 58.3 | 145.9 | 243.1 | 37.3 | 85.8 | 181.9 | 299.5 | 440.9 | 613.3 | 756.3 | 862.0 |
| Upper GI | 0.6 | 1.1 | 2.0 | 40.8 | 87.5 | 179.9 | 336.5 | 545.6 | 828.6 | 1204.9 | 1594.0 | 1973.2 |
| Urinary tract | 0.6 | 1.4 | 3.9 | 8.0 | 14.8 | 93.4 | 169.2 | 299.0 | 474.6 | 656.3 | 839.5 | 1071.2 |
| **MSH2** |  |  |  |  |  |  |  |  |  |  |  |  |
| Colon | 0.7 | 223.9 | 315.5 | 437.6 | 712.3 | 1322.9 | 2615.3 | 698.9 | 1172.7 | 1765.7 | 2392.5 | 3178.2 |
| Rectum | 0.3 | 0.6 | 93.1 | 155.1 | 291.6 | 670.1 | 199.0 | 327.6 | 482.2 | 670.8 | 827.2 | 942.7 |
| Upper GI | 0.6 | 1.1 | 2.0 | 4.2 | 9.0 | 193.0 | 361.0 | 585.3 | 888.8 | 1292.6 | 1709.9 | 2116.7 |
| Urinary tract | 0.6 | 1.4 | 3.9 | 8.0 | 221.2 | 390.2 | 707.1 | 1249.7 | 1983.7 | 720.7 | 921.7 | 1176.2 |
| **MSH6** |  |  |  |  |  |  |  |  |  |  |  |  |
| Colon | 0.7 | 2.2 | 34.4 | 47.8 | 77.7 | 144.4 | 285.4 | 527.6 | 885.2 | 1332.8 | 1806.0 | 2399.1 |
| Rectum | 0.3 | 0.6 | 1.5 | 2.5 | 4.7 | 10.8 | 22.9 | 37.7 | 256.8 | 357.3 | 440.6 | 502.1 |
| Upper GI | 0.6 | 1.1 | 2.0 | 4.2 | 9.0 | 18.5 | 100.4 | 162.7 | 247.1 | 359.4 | 475.4 | 588.6 |
| Urinary tract | 0.6 | 1.4 | 3.9 | 8.0 | 14.8 | 64.0 | 115.9 | 204.9 | 325.3 | 449.8 | 575.3 | 734.1 |
| **PMS2** |  |  |  |  |  |  |  |  |  |  |  |  |
| Colon | 0.7 | 2.2 | 3.1 | 4.3 | 7.0 | 13.0 | 25.7 | 47.5 | 79.7 | 1287.1 | 1744.0 | 2316.8 |
| Rectum | 0.3 | 0.6 | 1.5 | 2.5 | 4.7 | 10.8 | 22.9 | 37.7 | 104.2 | 144.9 | 178.7 | 203.7 |
| Upper GI | 0.6 | 1.1 | 2.0 | 4.2 | 9.0 | 28.5 | 53.2 | 86.3 | 131.1 | 190.6 | 252.1 | 312.1 |
| Urinary tract | 0.6 | 1.4 | 3.9 | 8.0 | 14.8 | 26.1 | 47.3 | 83.6 | 132.7 | 183.5 | 234.7 | 30.0 |

**Table 2:**
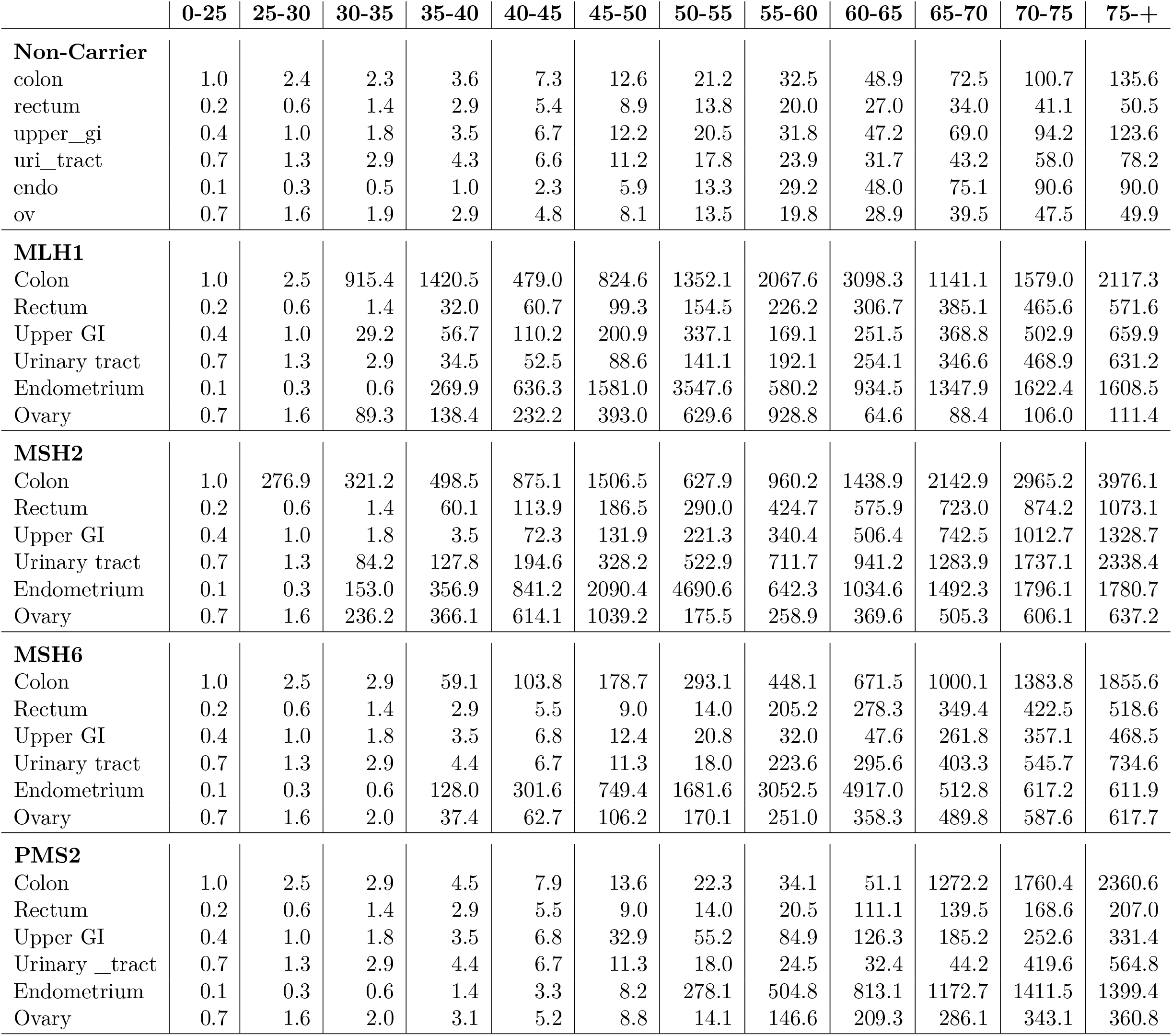
Incidence rate estimates in females per 100,000 person-years. Same layout and definitions as in Table S1.

MMR PV frequencies are half that of heterozygous carriers estimated by Win et al. ^14^ at *q* = (1*/*1946, 1*/*2841, 1*/*758, 1*/*714) for (MLH1, MSH2, MSH6, PMS2) PV carriers. Diagnostic performance of somatic tests are those reported by Assasi et al. ^15^ at (0.900, 0.947, 0.820, 0.570) for the sensitivity and (0.810, 0.770, 0.960, 0.980) for the specificity of (IHC, MSI testing, MLH1 promoter methylation analysis, BRAF testing).

### Computation

The main formula of the model writes

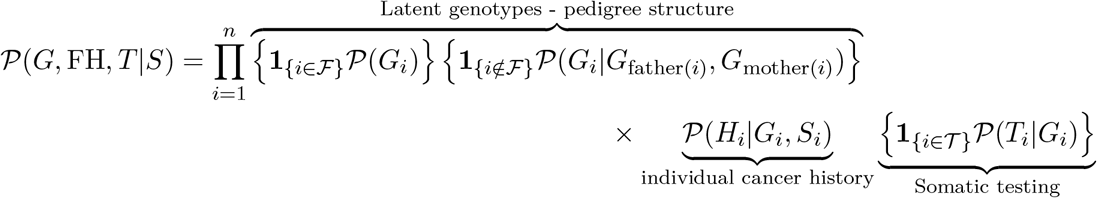

for a family composed of *n* members where *F* denotes for the set of founders and *T*, the set of family members who received somatic testing. *G* = (*G*_1_, …, *G*_*n*_) is the vector of individual genotypes, FH = (*H*_1_, …, *H*_*n*_) is the vector of individual cancer histories (including all family members, wether diseased or not), *T* = (*T*_*i*_)_*i*∈*T*_ is the vector of somatic testing and *S* = (*S*_1_, …, *S*_*n*_) is the vector of individual sexes.

The individual contribution of a cancer history is expressed in a competing risk setting where the censoring is independent from cancer. Moreover, somatic test results are assumed to be conditionally independent given the genotype, with the exception of (i) MSI and MMR IHC, (ii) MLH1 promoter hypermethylation and BRAF mutation testing, and (iii) MLH1 promoter hypermethylation, BRAF mutation testing, and MMR IHC. Moreover, in the model, loss of MLH1/PMS2 (or MSH2/MSH6) dimer by IHC contributes evidence for MLH1 (or MSH2) only, while being considered uninformative for PMS2 (or MSH6) with respect to LS status.

The posterior probability of the genotype of any family member is obtained by direct application of Bayes formula

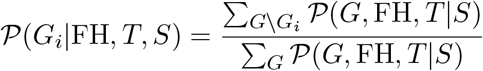

where 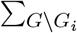 stands for the sum over all possible values of all genotypes of family members except the genotype of Individual *i*. We use classical message-passing algorithms applied to pedigree analyses for the inferences ^16;17^.

In the following, a posterior probability of LS is expressed as a percentage and referred to as a LynchRisk score (LRS).

## Results

To illustrate the potential clinical utility of LynchRisk, we present two simulated cases. The age at which LRS was calculated was the age at cancer diagnosis.

### Case 1

This female proband was diagnosed with MMRp (IHC) colon cancer at age 51 (figure 1a). LS seems unlikely given the MMRp status and the age at diagnosis. Indeed, LRS is only 0.14% in the absence of Lynch-associated cancer FH (figure 1b). However, FH has a major impact on LRS, with potential implications regarding germline testing. Indeed, LRS increases to 2.85% if the grandmother had endometrial cancer at age 45. It rises even further - 10.23% - if the proband’s mother is uninformative, for example if she died of an accident aged 28. One would definitely want to prescribe germline testing with LRS of 10.23%, while the decision at a 2.85% estimate would depend on local practice and available resources.

**Figure 1:**
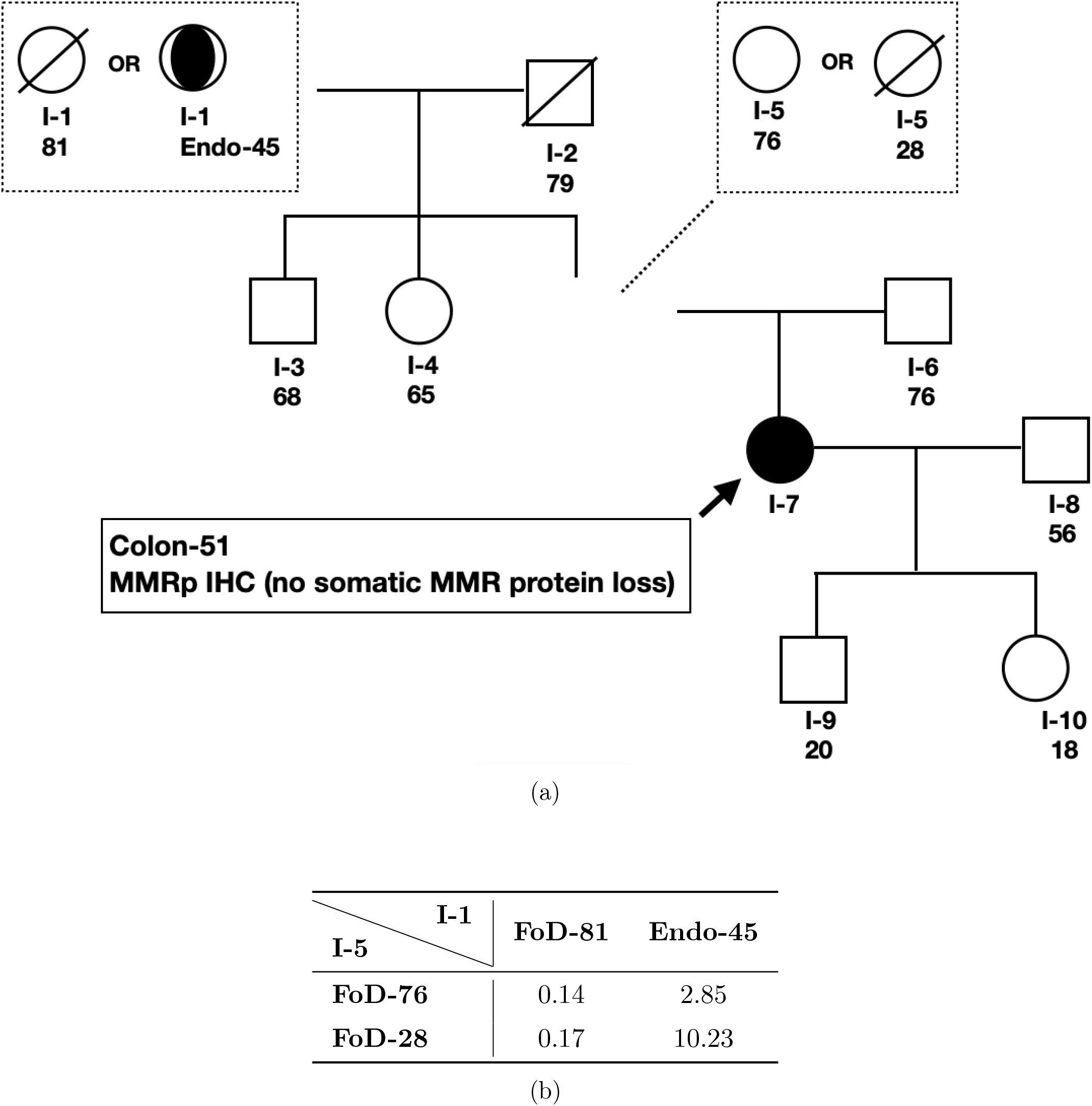
**Case 1. (a)** The female proband has a diagnosis of MMRp colon cancer aged 51, with different family histories: grandmother unaffected or with endometrial cancer aged 45, and mother unaffected aged 76 or deceased aged 28 of an unrelated cause. **(b)** Associated LynchRisk Scores (LRS) as the probability of Lynch syndrome expressed in percent (%). FoD stands for Free of Disease.

If the proband’s cancer was MMRd (MLH1/PMS2 loss with no somatic MLH1 promoter hypermethylation), LRS would be 7.38% and 9.01% in the absence of FH, but would rise to 72.28% and 92.09% with a grandmother diagnosed with endometrial cancer aged 45 without and with a mother deceased of a non-cancer cause at the age of 28, respectively. Germline testing would be warranted regardless.

### Case 2

Case 2 initially appears more suggestive of LS than Case 1. This male proband had a diagnosis of colon cancer aged 64. The cancer was MMRd, with MLH1/PMS2 loss and no somatic MLH1 promoter hypermethylation (figure 2a). In the absence of LS-associated cancer FH, LRS is only 0.49% (figure 2b), questioning the need for germline testing, regardless of MMRd. It decreases further to 0.24% when two unaffected sisters are added. However, LRS rises to 9.88% when a colon cancer at age 51 in the father is added.

**Figure 2:**
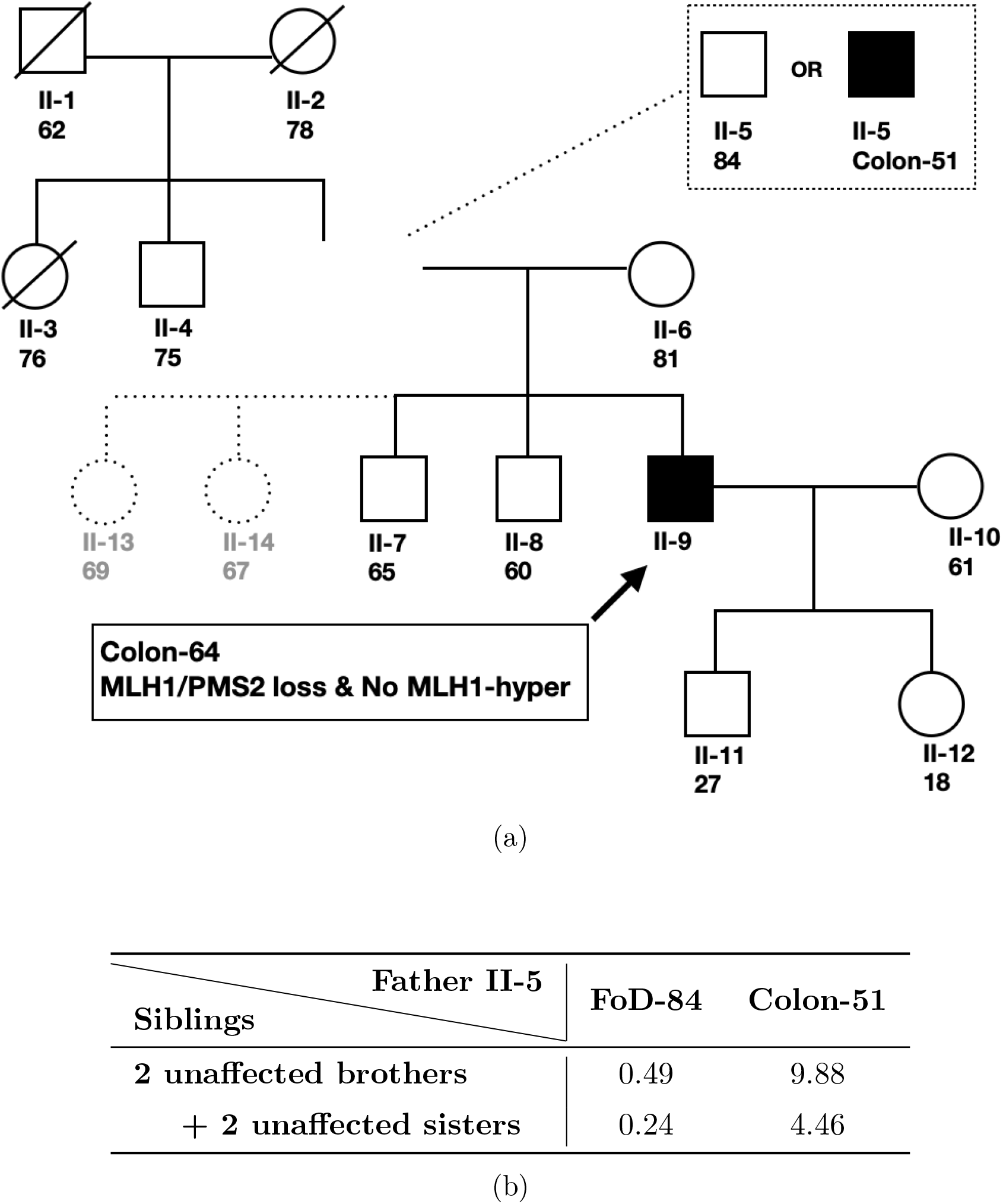
**Case 2. (a)** The male proband had a diagnosis of MMRd colon cancer aged 64, with different family histories: father unaffected or with colon cancer aged 51, two unaffected brothers or four unaffected siblings (two brothers, two sisters) **(b)** Associated LynchRisk Scores (LRS) as the probability of Lynch syndrome expressed in percent (%).

These contrasting LRS estimates could have clinical implications beyond germline testing. Assuming testing is indeed carried out on the basis of the MMRd colon cancer, negative results might be considered conclusive with an LRS of 0.49% (i.e. confidently rule out LS), while further investigations, such as long-read sequencing and somatic MMR sequencing would be considered in the second scenario (LRS of 9.88%).

Again the MMR status has a major impact on LRS. With an MMRp colon cancer in the proband, LRS is only 0.46% with a history of colon cancer aged 51 in the father, decreasing to 0.23% if two unaffected sisters are added.

## Discussion

We developed LynchRisk, a Bayesian framework that estimates LS probability in patients. Its strengths is the integration of family structure – including unaffected relatives -, personal and family cancer history, and MMR status into a unified prediction model.

The two cases presented in the results show how much detailed family structure and MMR status influence probability estimates. In Case 1, a female with MMRp colon cancer at the age of 51 and a FH of endometrial cancer, LRS increased from 2.85% to 10.23% if her mother was uninformative (e.g. deceased at a young age of a non-cancer cause). Germline testing might not be systematic at the 2.85% estimate in countries with limited resources, while it would certainly be prescribed at >10%. Case 2 was a male proband with colon cancer aged 64, and with a colon cancer history in the father at the age of 51. LRS was 9.88% if his cancer was MMRd, but dropped to 0.46% should the cancer be MMRp. Again, such different estimates would likely have an impact on germline testing decision.

LynchRisk could also provide valuable information beyond LS germline testing. One often encounters negative germline testing in a patient with an MMRd cancer. There are two possible explanations for this: (a) the association of two somatic MMR genetic events^18^, or alternatively (b) the existence of a complex germline variant that escapes detection by standard next-generation sequencing ^19^. Neither somatic nor long-read germline sequencing are universally available outside of high-volume high-resource centers, and LRS could prioritize patients for further investigations in this context.

LS mathematical models have been developed in the past 20 years. The most widely used, PREMM5, has excellent sensitivity and predictive value at a 2.5% probability threshold^20^. However, it is based exclusively on Lynch-associated cancers in the patient or his relatives. It does not account for unaffected relatives nor does it consider MMR deficiency of proficiency. We have shown with LynchRisk how these parameters influence LS probability. Furthermore, the growing availability of clinical and tumour data facilitates their integration into clinical models, particularly as MMR status is increasingly explored given its therapeutic implications.

Several limitations should be acknowledged. LynchRisk relies on published MMR PV prevalence, cancer incidence data and somatic MMR test performances, which may evolve over time as ever larger studies are published. Piecewise-constant hazard functions were adopted to ensure computational tractability; they may induce abrupt changes in the contribution of a given cancer history at specific ages. Finally, it has not been validated in patient cohorts.

In conclusion, LynchRisk provides an interpretable framework for integrating pedigree, cancer history, and tumour characteristics into Lynch syndrome probability estimates. Further work will focus on recalibrating the cancer hazard functions and validating model performance through simulations. We also plan to develop an R package to make LynchRisk available to research groups interested in its evaluation.

## Data Availability

All data produced in this manuscript are simulated and described within the manuscript.

## Additional information

## Acknowledgments

The authors warmly thank La Ligue contre le cancer who funded this work (PhD grant). We would like to thank Florence Coulet, Erell Guillerm, and Alex Duval for fruitful discussions on Lynch syndrome. We also thank Alex Duval for his help in securing funding. We are grateful to Dominique Stoppa-Lyonnet and Antoine de Pauw for sharing their insight on hereditary cancer syndromes.

## Authors’ contributions

AL: Conceptualization, Formal analysis, Investigation, Methodology, Software, Validation, Visualization, Writing - Original draft, Writing - Review & editing. PB: Conceptualization, Investigation, Methodology, Project administration, Supervision, Visualization, Writing - review & editing. GN: Conceptualization, Investigation, Methodology, Project administration, Software, Supervision, Writing - Review & editing.

## Appendix

### A Supplementary methods: calibration of cancer incidence rates

Hazard functions are assumed to be piecewise constant by time intervals of 5 years between age 25 and age 75. They are parametrised by the set of vectors of age-, sex-, cancer site-, and genotype-dependent incidence rates denoted *α*. At any age, the hazard function for cancer *k* conditional on genotype *g* and sex *s* denoted 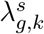 takes the value of *α* in related cancer site, genotype, sex and age interval.

LynchRisk considers six cancer sites: colon, rectum, endometrium, ovary, upper gastrointestinal tract (which aggregates stomach, small bowel, bile duct, gall-bladder, and pancreas) and urinary tract (which aggregates ureter, renal pelvis, and urinary bladder).

### Calibration of the vectors of incidence rates in carriers of one mismatch repair (MMR) pathogenic variant (PV)

In order to reduce overfitting issues, the calibration of *α* started with a smoothing process of cancer incidence rates in carriers of a single MMR PV using cancer incidence rates in the French general population (GP) estimated by Defossez et al. ^11^ and incidence data (number of cancer cases and person-years) in carriers of one MMR PV of the Prospective Lynch Syndrome Database (PLSD, https://plsd.eu) reported by Dominguez-Valentin et al. ^12^ in their Table S5 (see Supplementary Information files of the authors).

Dominguez-Valentin et al. ^12^ provided cancer-specific incidence data under a non-competing-risk framework. For each cancer site, (i) observation time was censored at the occurrence of the first cancer in that site, while (ii) a history of cancer in other organs was disregarded. We assume that the incidence estimates reported by Defossez et al. ^11^ follow the same principles, with rare exceptions to assumption (ii) arising from specific histological profiles used to define multiple primary cancers. Accordingly, when considering a competing-risk framework, we define the cause-specific hazard for an aggregated cancer site as the sum of the cause-specific hazards of the corresponding individual localizations.

For any genotype *g* composed of a single MMR PV and *t >* 0, we assume that

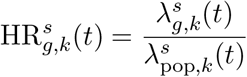

where 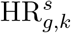 is the piecewise constant age-, sex- and cancer site-dependent hazard ratio between carriers of genotype *g* and the GP and 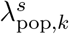 (*t*) is the incidence rate of cancer *k* in the GP at related age.

Under a piecewise constant hazard (PCH) model, the log-likelihood of log-hazard ratios can be expressed in terms of number of events and person-years at risk^21^. Log-hazard ratios were then smoothed by model selection using a greedy backward stepwise selection algorithm based on a likelihood ratio test with a p-value threshold of 1%.

### Calibration of the vectors of incidence rates in non-carriers

In the framework of a PCH model where the vector of cuts is denoted (*c*_0_, …, *c*_*j*_, …), the incidence rate for cancer *k* in non-carriers in time interval [*c*_*j*−1_, *c*_*j*_) is estimated by

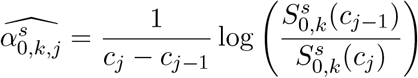

where 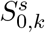 is the survival function for the time to first diagnosis of cancer *k* in non-carriers.

Survival functions 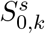 were previously calibrated using the expression of a finite mixture model in survival analysis^22^ under the assumption that the GP is solely composed of non-carriers and carriers of a single MMR PV. Therefore we can write

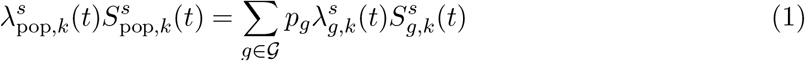

where *G* is the set of genotypes composed of zero or one MMR PV, *p*_*g*_ is the frequency of genotype *g* ∈ *G*, 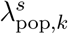 are respectively the survival and hazard functions for the time to first diagnosis of cancer *k* in the GP, 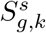 and 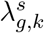 are those in carriers of genotype *g* ∈ *G*. We used genotype frequencies estimated by Win et al. ^14^ for the calibration. Moreover, survival functions of Equation 1 related to the GP and LS carriers of a single MMR PV are simply given, for *h* ∈ {*G \* {0} ∪ pop} by

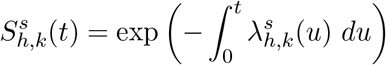

and computed using closed form expressions in PCH models.

Calibrated incidence rates in non-LS-carriers and carriers of a single MMR PV are reported, per 100,000 person-years, in Tables 1 and 2 for males and females respectively.

### Incidence rates in carriers of two MMR PVs

Finally we assume an additive effect of MMR PVs on the hazard scale for carriers of two PVs (in different MMR genes).

